# Detecting Heart Failure using novel bio-signals and a Knowledge Enhanced Neural Network

**DOI:** 10.1101/2022.10.26.22281541

**Authors:** Marta Afonso Nogueira, Simone Calcagno, Niall Campbell, Azfar Zaman, Georgios Koulaouzidis, Anwar Jalil, Firdous Alam, Tatjana Stankovic, Erzsebet Szabo, Aniko B. Szabo, Istvan Kecskes

## Abstract

1

**Background:** Clinical decisions about Heart Failure (HF) are frequently based on measurements of left ventricular ejection fraction (LVEF), relying mainly on echocardiography measurements for evaluating structural and functional abnormalities of heart disease. As echocardiography is not available in primary care, this means that HF cannot be detected on initial patient presentation. Instead, physicians in primary care must rely on a clinical diagnosis that can take weeks, even months of costly testing and clinical visits. As a result, the opportunity for early detection of HF is lost.

**Methods and results:** The standard 12-Lead ECG provides only limited diagnostic evidence for many common heart problems. ECG findings typically show low sensitivity for structural heart abnormalities and low specificity for function abnormalities, e.g., systolic dysfunction. As a result, structural and functional heart abnormalities are typically diagnosed by echocardiography in secondary care, effectively creating a diagnostic gap between primary and secondary care. This diagnostic gap was successfully reduced by an AI solution, the Cardio-HART™ (CHART), which uses Knowledge-enhanced Neural Networks to process novel bio-signals. Cardio-HART reached higher performance in prediction of HF when compared to the best ECG-based criteria: sensitivity increased from 53.5% to 82.8%, specificity from 85.1% to 86.9%, positive predictive value from 57.1% to 70.0%, the F-score from 56.4% to 72.2%, and area under curve from 0.79 to 0.91. The sensitivity of the HF-indicated findings is doubled by the AI compared to the best rule-based ECG-findings with a similar specificity level: from 38.6% to 71%.

**Conclusion:** Using an AI solution to process ECG and novel bio-signals, the CHART algorithms are able to predict structural, functional, and valve abnormalities, effectively reducing this diagnostic gap, thereby allowing for the early detection of most common heart diseases and HF in primary care.

## 2 Introduction

Cardiovascular disease (CVD) remains the leading cause of disease burden in the world, with 18.6 million CVD deaths registered in 2019 [1]. Heart Failure (HF) affects a large spectrum of this population, occurring at all ages and equally in both genders, with 38 million people affected globally [2]. HF increases with age, high blood pressure, obesity, and diabetes. Early detection is critical to accessing appropriate and timely treatment to achieve the best possible outcomes [3]. Nearly 80% of all HF diagnoses are initially made in emergency care [4], and this is despite over 60% of these same patients presenting to primary care with signs and symptoms of HF in the 6 months before their diagnosis. Delay and missed diagnoses are common, typically confirmed only after symptoms have become obvious and severe damage has set in. Estimates for delays for a HFrEF diagnosis ranges from several months in Germany to up to a year in Ireland, with serious delays in France, Netherlands, Sweden, and the UK [5]. For HFpEF, in an ongoing study of AF in the UK, it was reported as 12+ months [6].

ECG and natriuretic peptide testing play a role in the clinical decision leading to a HF diagnosis, but they have limitations that contribute to high false positive and low detection rates in the early phase. Access to echocardiography is all too often delayed due to backlogs and long waiting lists.

HF is characterized mainly by echocardiography measurements, and to a much lesser extent, ECG measurements, that can identify functional and structural heart abnormalities [7], which underpin the HF condition. LVEF, LV global longitudinal strain (GLS) [7][8][9] or LV volume or stroke volume (SV) [8] could be useful for early detection of HF, but these echocardiography measurements are only available in secondary care, not in primary care. Due to the lack of a non-invasive medical device for use in primary care able to provide echocardiography equivalent findings, the opportunity for early detection of HF is missed.

This research analyses the use of novel bio-signals enhanced with AI, to provide echocardiography equivalent findings for use in primary care settings, enabling the early detection of HF. Such bio-signals can provide a more precise estimation of structural and functional cardiac abnormalities that include LVEF, GLS, SV, LV mass index (LVMI), and left atrial volume index (LAVI) parameters for detecting and predicting HF.

A breakthrough technology from Cardio-Phoenix™, Cardio-HART™, based on complementing and augmenting ECG and PCG with a novel bio-signal of a physiological nature, is analysed in this paper. It is a low-cost, non-invasive, cardiac assessment system for use in patient care settings, for the early detection of CVD, including myocardial infarction, functional and valve abnormalities, and HF, including its phenotype, HFpEF, HFmrEF, or HFrEF.

### 2.1 Characteristic Nature of Heart Failure

HF is mainly categorized based on LVEF, including HFpEF (50≤LVEF), HFmrEF (50>LVEF>40), and HFrEF (40≥LVEF) as confirmed by the recent guidelines [3][10]. However, HF diagnostic rules and algorithms include structural heart disease criteria according to guidelines [3][10][11]. To achieve a clinically meaningful outcome, each HF category should be characterized by its characteristic dysfunctions, for which LVEF has limitations [12]. LVEF alone is insufficiently sensitive for subtle LV systolic dysfunction caused by regional wall motion abnormality (WMA), which is better detected by mitral annular systolic excursion or systolic velocities or LV global longitudinal strain (GLS) [7][8][9]; LVEF correlates less with long-axis functional reserve of the LV or with peripheral blood flow. Still, LVEF, GLS, or SV could be useful for the early detection of HF with the extension of structural criteria [13].

#### 2.1.1 HFpEF characteristics

HFpEF should be suspected in patients with typical HF symptoms and signs (S3 heart sound, displaced apical pulse, and jugular venous distension) of chronic heart failure [14].

HFpEF is suspected when an echocardiography finding of a normal LVEF is paired with relevant structural heart disease or diastolic dysfunction (DD) [3][10][14]. However, HFpEF cannot be diagnosed from a single echocardiographic parameter, but inclusion of functional and structural parameters (AF, E/e’, RVSP, GLS, LAVI, LVMI) with risk factors (Elder Age, Hypertensive, Obesity) together with a natriuretic peptide test can better define this heterogeneous disorder [11][15]. Additionally, HFpEF patients are also likely to have QRS, QTc, and PR interval prolongation or pacemaker on ECG, compared to non-HF patients [15].

#### 2.1.2 HFmrEF characteristics

Patients with HFmrEF have a different clinical profile, but more similar to patients with HFpEF [16], primarily mild LVSD, but with features of DD. The diagnostic criteria for HFmrEF include any relevant structural heart disease, such as LVH or LAE or DD besides symptoms and the mildly reduced EF (LVEF of 40-49%) [10]. HFmrEF patient’s typical structural abnormality is mild DCM, the typical functional abnormality is WMA with either MI or ST-T deviation, and the typical electrophysiological problem is AFib, PAC, and lower T wave axis (valve diseases are less typical when compared to HFpEF) [13].

#### 2.1.3 HFrEF characteristics

HFrEF is most commonly associated with DCM or CAD [17]. LVSD and moderate AS commonly occur together; patients with moderate AS and concomitant LVSD are at high risk for clinical events including all-cause death, hospitalization for HF, and aortic valve replacement [18]. HFrEF group of patients have the highest levels of comorbidity and severity: the typical structural abnormalities are the DCM, left and right atrial and ventricular enlargement; the typical functional abnormality is LVSD, the typical hemodynamical problem is mitral/tricuspid regurgitation and PH, and all the common ECG abnormalities show higher prevalence, including LVH and RVH criteria [13].

### 2.2 Bio-Signal Characteristics

Diagnosing HF related abnormalities using synchronized bio-signals depends on the prediction capability of the underlying echocardiographic characteristics associated with HF. This article demonstrates that other non-invasive bio-signals, phonocardiography, and mechano-physiological, can be used to augment and complement ECG in such a way that they can provide comparable accuracy and reproducibility estimation of LV systolic and diastolic function, regarding echocardiography, with clinically acceptable diagnostic power.

The bio-signal capabilities to assess systolic dysfunction and typical signs and comorbidities of HF are discussed in the following subsections.

#### 2.2.1 Electrocardiogram

ECG interpretation can reach high performance if the condition is related to the electrophysiology of the heart, such as specific arrhythmias, premature beats, atrioventricular blocks, and bundle-branch blocks. But ECG is less clear in the case of mechano-physiological or hemodynamical abnormalities.

ECG has some signs for the mechano-physiological diseases, typically the repolarization abnormalities, (abnormal ST-T) associated to CAD, the abnormally high QRS voltages as a manifestation of ventricular hypertrophy and abnormal P-wave voltages related to atrial enlargement.

The standard reading of ECG shows low sensitivity (20-50%) with moderate-high specificity (85-90%) for ventricular hypertrophy [19][20] and atrial enlargement [21][22], where these structural abnormalities are defined by standard LVMI, IVSd, RWT, LAVI, and RAVI echocardiographic parameters. In practice, this results in high levels of FN.

Some recent ECG processing, shows the prediction capability for severe systolic dysfunction and moderate/severe aortic stenosis (AS) [23], but the results demonstrate that the ECG signs (essentially inverted T-waves) are not sufficiently specific for any disease and the PPV remains low, 10-30%, again leading to higher levels of FP. ECG signals processed with state-of-the-art NNs can arguably reach workable sensitivity for patients having LVEF<35% [24]. However, the positive predictive value (PPV) is typically low (10-30%), because ECG signs for low EF do not have sufficient specificity.

#### 2.2.2 Phonocardiography

Phonocardiography (PCG) signals capture heart sounds that contain useful and familiar information about the condition of the heart for the early detection and diagnosis of some heart diseases [25], in particular, valve diseases [26] and HF [27]. The acoustic waveform of PCG caused by the movement of blood in the cardiovascular system is considered an important indicator in the diagnosis of several CVDs [28] and hemodynamic findings, such as Atrial Stenosis, or “AS” [29]. Advanced electronic stethoscopes can detect heart murmurs [26] with higher performance, including some valve diseases, and with moderate performance for mitral and aortic regurgitation [30].

#### 2.2.3 Mechano-physiological force

Bio-signal measuring mechanical physiological force correlates to cardiac activity [31], which has similar measurement characteristics to GLS and SV [32][33], because it can capture both wall motion at four different auscultation points and myocardial contractility. The characteristics of these signals contain useful information that correlates with cardiovascular physiological and pathological processes [34] and dysfunctions. This method can predict systolic and diastolic left ventricular function in ischemic heart disease, as indicated by basic research [35][36]. For example, one study resulted in a high correlation between the change in signal derived “g-value” and the change in EF% (r=0.87) [37].

Extensive research and development has resulted in a bio-signal that measures cardiac characteristics of a mechano-physiological nature (MCG). Together, when this novel bio-signal is synchronized and combined with ECG and PCG bio-signals, they can provide sufficient characteristic information about cardiac dysfunctions associated with various CVDs, including HF. However, the extraction of cardiac dysfunction from these signals requires effective processing techniques by an advanced AI system.

#### 2.2.4 Systolic Time Intervals

Systolic time intervals are an alternative bio-signal-based evaluation of abnormal LV systolic function [38], which can be measured by synchronized bio-signals of an electrical, hemodynamic, and physiological nature: systolic ejection time (SET) [39], systolic performance index (PEP/LVET) [40], and electromechanical activation time (EMAT) [41].

Research pairing bio-signals with ECG has been shown to predict some cardiovascular diseases [35], e.g., systolic and diastolic function in ischemic heart disease [36]. The measurement of PEP and LVET is a common point between ECHO and bio-signals, and a study resulted in a 0.6-0.8 correlation between ECHO-based and bio-signal based PEP measurements [42].

### 2.3 Organization of the Article

In the introduction, we share background knowledge about how the bio-signals are able to detect heart failure. In methods, a brief summary of the Cardio-HART algorithm is provided, focusing on its unique modeling. Then, in the results, the performance of the Cardio-HART solution is presented and compared to a traditional ECG. The discussion looks at the implications of the solution in health care systems and is followed by a conclusion.

## 3 Methods

The CHART system captures and its AI processes three types of bio-signals. Each bio-signal has its own strengths:

- ECG – Electro-physiological abnormalities: arrhythmias, premature beats, atrioventricular blocks, bundle-branch blocks, etc.
- MCG – Mechano-physiological abnormalities: hypertrophy, atrial enlargement, systolic or diastolic dysfunction, cardiomyopathy, myocarditis, myocardial infarction, ischemia, or other wall motion problems
- PCG - Hemodynamic diseases: valve stenosis, valve regurgitation, hypertension (arterial or pulmonary hypertension)

CHART’s AI, synchronously combines and processes the ECG, PCG, and MCG bio-signals to predict, with high probability, LV systolic and diastolic dysfunctions, as well as structural and valve related abnormalities connected to HF, similarly to ECHO in the form of HART-findings.

HART-findings™ are disease equivalent to ECHO-findings, but derived using bio-signals rather than from images as in Echocardiography^1^.

As abnormalities and comorbidities of HF are essentially diagnosed primarily by ECHO and to a much lesser extent by ECG, the classification of HF can be commonly resolved by CHART’s AI.

As ECHO is only available in secondary care, the prediction of HF lacks an appropriate device for use in primary care.

### 3.1 Cardio-HART Algorithm

Cardio-HART is an AI-powered, cardiac diagnostic system for use in clinical care settings, including primary and secondary care. It starts in a clinical care setting where the CHART device first captures the bio-signals, ECG, PCG, and MCG, and then uploads them to the cloud for AI processing. The CHART AI then outputs a wide range of medical findings or endpoints, consistent with cardiac dysfunction.

As a result, Cardio-HART has the diagnostic functionality characteristic of three different cardiac devices combined, including: 1) 12-lead ECG; 2) eStethoscope (Phonocardiography); and 3) Echocardiography.

Uniquely, Cardio-HART can diagnose 14 HART-findings, typically only diagnosed by echocardiography in cardiology. The HART-findings are disease equivalent to ECHO-findings. They are classified as “Normal/Mild/Abnormal” for the 14 common abnormalities listed in Table I, including: structural problems (LVH, DCM, RVE, LAE, RAE), functional problems (WMA, LVSD, DDIM), and valve problems (AS, MS, AR, MR, TR, PH).

From these HART findings, HF is then classified into 4 phenotypes, consistent with their medical context: HF Unlikely, HFpEF, HFmrEF, and HFrEF.

#### 3.1.1 Cardio-HART AI

Cardio-HART AI is an extensive set of interconnected and related algorithms that prepare, process, and analyze the medical information (see Fig. 1), to extract both signal-based findings and machine learning findings that together contribute to the final outputs related to heart disease and cardiac status, including:

1. **Signal preprocessing**: cleaning the bio-signals using digital filters, artifact removal, and 3D stabilization techniques.
2. **Signal segmentation** – Heart cycles are segmented into characteristics points and into typical beats, taking advantage of synchronized bio-signals. The ECG is segmented according to standards: P- Q-, R-, R’-, S-, S’-, T-wave. Heart sounds segmented on PCG signals as: S1-onset, S1-peak, S1-offset, S2-onset, S2-peak, S2-offset. Aortic opening AO and closure AC, mitral opening MO and closure MC are detected on the MCG signals.
3. **Signal measurements** – Extracts measurement from the segmented bio-signals including amplitudes, intervals, time-frequency representation, cross-power spectrum density and statistical features. The important systolic time intervals can be calculated thanks to the synchronized and segmented bio-signals, such as PQ-, QRS-, QT-interval, Q-S1, Q-S2 intervals, EMAT, PEP, LVET, etc.
4. **Signal-based findings:** Medically important abnormalities are detected based on known medical rules. PCG sound amplitude measurements use semi-supervised parameters, e.g. for systolic murmur. It uses dynamic thresholds in function of patient characteristics, e.g. prolonged PQ interval using gender and heart rate normalized thresholding.
5. **Signal Transformation**: This prepares the processed signals for the neural networks using digital signal processing, segmentation-based alignment, and normalization. It reduces the number of network layers and, indirectly, the required number of samples, which is key in avoiding overlearning. Three main transformations are applied:
  1) typical beat averaging, which strongly reduces noise;
  2) aligned beat sequence (21 beats), which preserves beat variability;
  3) time-frequency representation, which highlights the frequency domain of the signals.
6. **Feature compression** – This supervised process reduces the number of features to produce disease-specific features. State-of-the-art feature compression methods are then used in multi-layer convolution neural networks to extract the medically useful information from the complex or multi-dimensional data (bio-signal). See Fig. 3 for architecture details.
  a. ECG: Convolutional long-short-term memory (LSTM) and convolutional capsule networks are utilized for processing the transformed ECG signal and its softmax classification layer is trained for the reference ECG findings (e.g., MI presence/absence).
  b. HART: Same architecture networks are applied as at ECG, but used PCG and MCG signals too and trained for fitting 12 key echocardiographic measurements (LVEF, LVMI, LAVI, E/A, etc.). Regularization techniques are implemented to prevent overlearning, and Pearson correlation is calculated as the ultimate performance metric, which reaches 0.5-0.8 values on the validation datasets.
7. **Machine-learning-based findings**
  a. ECG: Knowledge-enhanced neural networks (KENN, similar to [52]) are used for classifying the complex multi-parameter^2^ ECG-findings (when the reference standard is not ECG), and the rest are rule-based according to ECG standards. KENN is a type of hybrid model, similarly to physics-enhanced neural networks, where the knowledge base rules are the primary model and NN (described in point 6) is the secondary or complementary model, which resolves borderline cases. The KENN successfully regularized the NN and the NN becomes active only in specific cases when the rule is uncertain. The decision is based on maximum probability, which comes, first, from sigmoid probability-based rules,

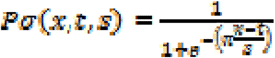

where *x* denotes measurements, *t* is the threshold of abnormal *x*, and *s* is a scale factor) and second, from NN softmax layer

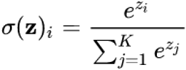

calculated for i=1..K classes, where *z* is the previous layer output.
  b. HART: Using both signal measurements and compressed features, the machine learning models address the echo-equivalence findings in triple-class format (Normal/Mild/Abnormal) as the top-level interpretation of heart failure and CVD – called HART-findings. Five feature selection methods are utilized, two are based on joint mutual information, one is based on Markov-blanket, one is eigenvector-based and one embedded in feature selection methods. Naïve Bayes, Discriminant Analysis, and Shallow Neural Network methods are used for classification. At the end of training, a wrapper-type feature selection method is used to prune the features. Ensemble learning selects the best combination from 5×3=15 models based on performance, overfitting, and expert supervision of selected features.
8. **HF indication –** It is a head layer on Cardio-HART-findings, which uses rules related to classifying HF. It is described in the next section of 3.2.
9. **Cardio-HART report generation:** Finally, the algorithm generates a colorful and multi-page report demonstrating how the segmented signals result in the measurements and findings. The report additionally provides HF classification with estimated LVEF and decision support for primary care users.

**Figure 1.**
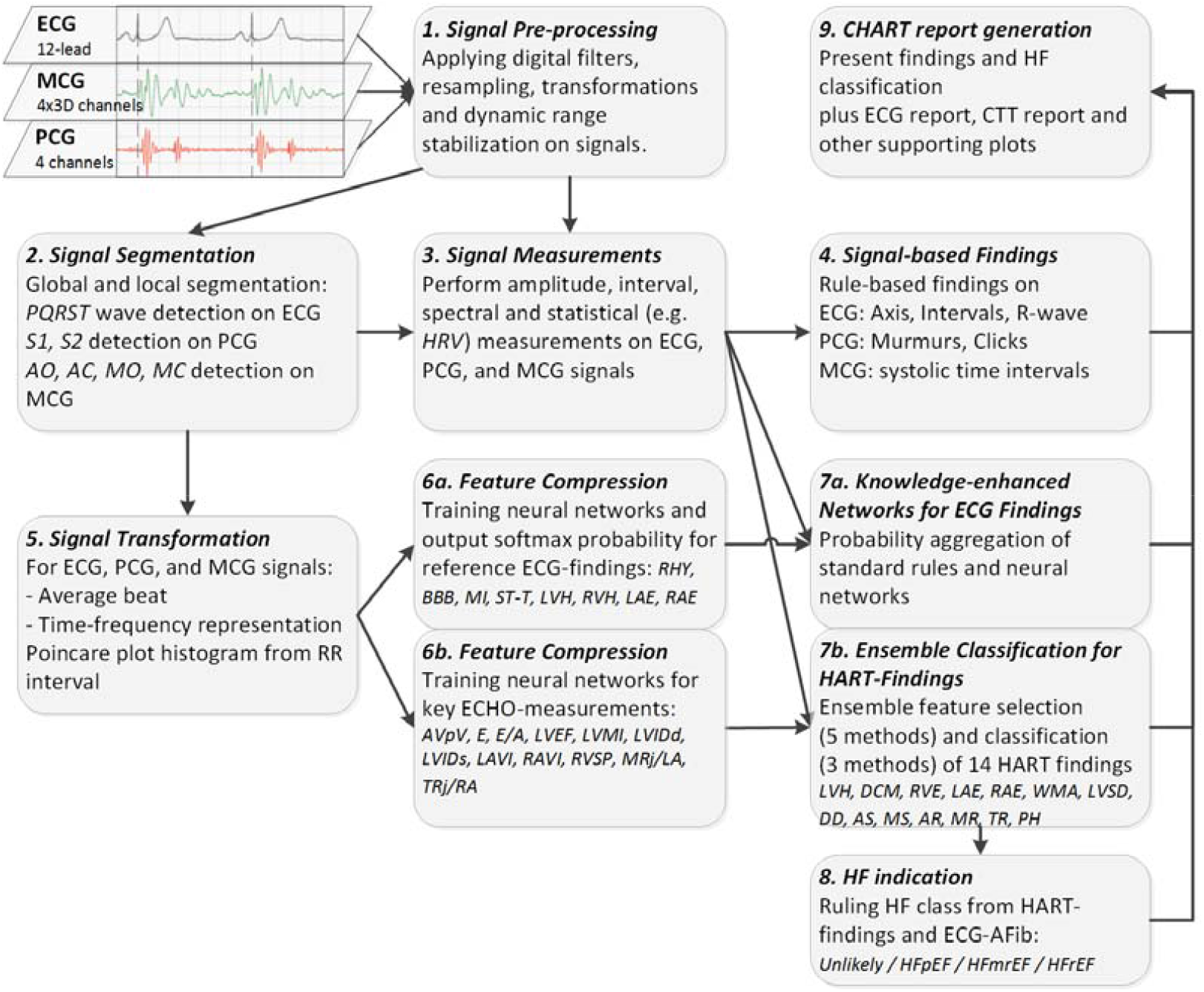
Main modules of Complete Cardio-HART algorithm that predicts HART-Findings and indicates HF.

Given its overall size, parts of the algorithm (e.g., points 1-4) are not detailed in this paper. More details can be found in the Cardio-HART whitepaper, available by request on the official website^3^.

#### 3.1.2 Training and Validation

Separate training and validation datasets were used. Training was provided using a unique training dataset of 14150 records, which was not used for performance validation. The neural networks split the training dataset into two, a training set and a test set. The ensemble feature selection and classification use a 5-fold cross-validation method on the training dataset to estimate performance. The finale classifiers are selected based on the maximum cross-validation performance and minimum estimated overfit. The overfit is estimated between training and cross-validation, but ultimately between cross-validation and external validation performance, see Fig. 2.

**Figure 2.**
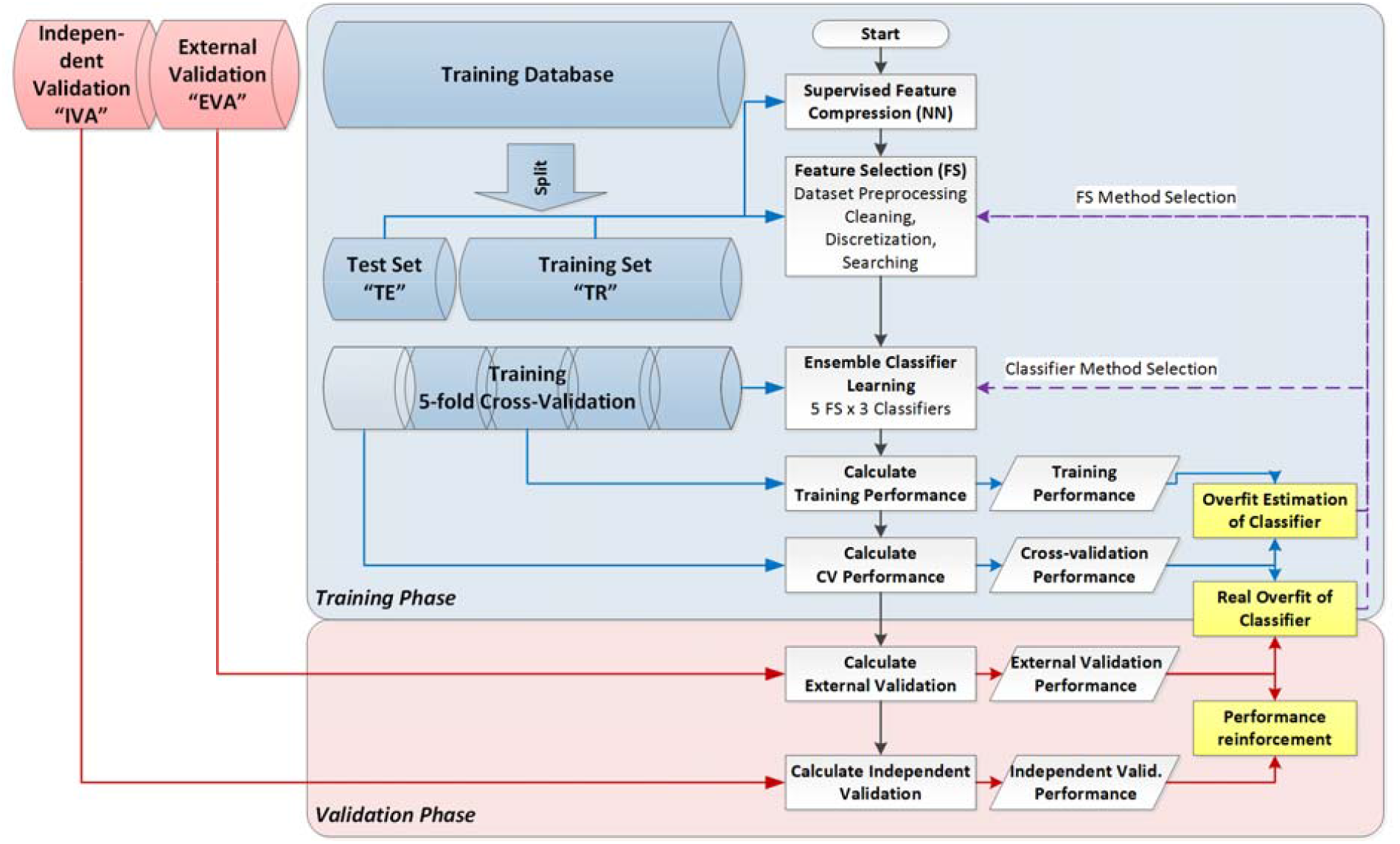
Splitting database for training and validation and checking overfit of classification.

### 3.2 HF indication model from HART-findings

The innovation analyses three types of bio-signals (mentioned in Table 1) to identify characteristics for each disease measured at four cardiac auscultation points. Using state-of-the-art neural network techniques such as Convolutional Recurrent Network (LSTM) and Convolutional Capsule Network (CapsNet), it extracts the hidden, synchronized and localized medical information from the signals. The supervised training of these networks is made for key echocardiographic measurements, including LVEF, as it detailed in point 6b. These compressed features provide the main prediction capability based on the estimated LVEF and LVSD findings and other HART-findings, HF categories are indicated using rules, as demonstrated on the bottom part of Fig. 3.

**Table 1.** External (EVA) and Independent (IVA) validation performance of 14 HART-findings by Cardio-HART.

| HART Finding | SE% |  | SP% |  | PPV% |  | NPV% |  | PREV% |  |
| --- | --- | --- | --- | --- | --- | --- | --- | --- | --- | --- |
|  | EVA | IVA | EVA | IVA | EVA | IVA | EVA | IVA | EVA | IVA |
| LVH | 80.9 | 77.2 | 81.1 | 95.7 | 60.6 | 77.9 | 92.2 | 95.5 | 26.5 | 16.5 |
| DCM | 78.1 | 70.7 | 90.4 | 98.2 | 61.5 | 81.9 | 95.4 | 96.7 | 16.4 | 10.2 |
| RVE | 67.1 | 69.7 | 94.9 | 97.9 | 49.4 | 44.2 | 97.5 | 99.3 | 6.9 | 2.3 |
| LAE | 97.2 | 95.9 | 92.4 | 97.5 | 64.7 | 73.8 | 99.6 | 99.7 | 12.5 | 6.7 |
| RAE | 85.9 | 61.8 | 92.6 | 96.9 | 50.4 | 50.0 | 98.7 | 98.1 | 8.0 | 4.7 |
| WMA | 87.4 | 81.8 | 90.0 | 95.6 | 49.8 | 69.9 | 98.4 | 97.7 | 10.2 | 11.0 |
| LVSD | 70.0 | 66.7 | 96.2 | 98.0 | 63.3 | 74.5 | 97.2 | 97.2 | 8.5 | 7.9 |
| DDIM | 74.8 | 77.5 | 80.4 | 78.6 | 61.3 | 72.8 | 88.5 | 82.5 | 29.3 | 42.5 |
| AS | 89.5 | 85.4 | 97.3 | 98.3 | 70.1 | 63.1 | 99.2 | 99.5 | 6.6 | 3.3 |
| MS | 64.9 | NA* | 98.0 | NA* | 48.5 | NA* | 99.0 | NA* | 2.8 | 0* |
| AR | 64.6 | 69.9 | 86.9 | 94.5 | 57.1 | 62.4 | 89.9 | 96.0 | 21.4 | 11.5 |
| MR | 70.3 | 83.3 | 84.6 | 97.2 | 60.7 | 70.9 | 89.4 | 98.6 | 25.3 | 7.5 |
| TR | 72.5 | 64.0 | 89.7 | 96.6 | 61.6 | 58.7 | 93.5 | 97.3 | 18.5 | 6.9 |
| PH | 67.0 | 79.5 | 95.9 | 99.0 | 52.4 | 71.4 | 97.8 | 99.4 | 6.2 | 3.1 |
| **Average** | **76.4** | **75.6** | **90.7** | **95.7** | **58.0** | **67.0** | **95.5** | **96.7** | **14.2** | **10.3** |

\*EV database has not sufficient positive for MS

**Figure 3.**
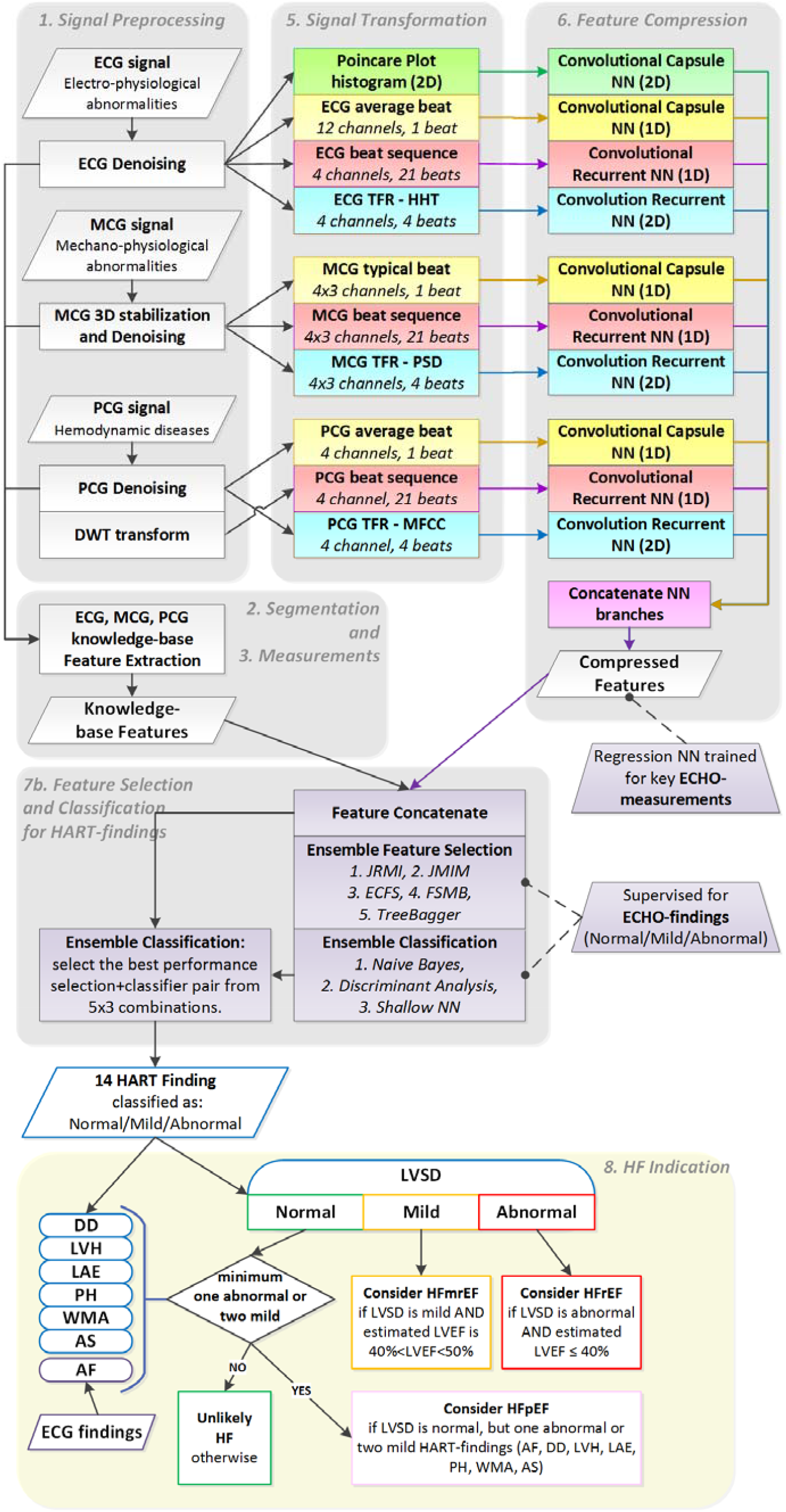
Cardio-HART block diagram for HART machine learning models and indicating HF.

### 3.3 Performance evaluation

The common binary performance metrics are calculated for the HART-findings validation, presented in section 4.1: sensitivity (*SE=TP/(TP+FN)*)^4^, specificity (*SP=TN/(TN+FP)*), positive and negative predictive value (*PPV=TP/(TP+FP), NPV=TN/(TN+FN)*), plus the prevalence is provided (*PREV=(TP+FP)/(TP+FP+TN+FN)*).

In the ECG versus HART comparison, we chose appropriate ECG performance metrics to make a fair comparison because ECG-findings have low specificity, typically for LVSD. The prevalence of ECG and ECHO-findings is imbalanced for the target population (typically 1-20%). Hence, the sensitivity (a.k.a. recall) is relevant, but the PPV (a.k.a. precision) is more representative compared to specificity. This is the reason for Precision-Recall Curve (PRC) analysis in addition to Receiver Operating Characteristic (ROC) analyses. The F-score is a combination of precision and recall and is an appropriate scalar performance metric for binary classification to choose the best predictive ECG-finding for ECHO-findings. The F-score is an appropriate scalar performance metric for binary classification to choose the best predictive ECG-finding for ECHO-findings as the F-score is a combination of precision and recall. Because of the imbalanced case (prevalence smaller than 50%), the beta was chosen to β=0.5 to regularize the too many false positives and ensure the prevalence-based balanced classification (denoted as “F05” on figures).

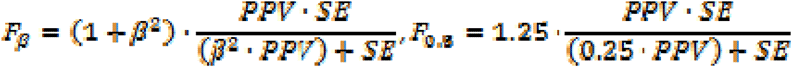

## 4 Results

### 4.1 Diagnostic Performance of HART-Findings™

The validation procedure of HART-findings is based on an ECHO-based ground truth. HART-findings were validated with a database having both parallel bio-signals and ECHO assessment. The performance shows sensitivity ranging between 65-90% with 90%+ specificity and 5-20% disease prevalence (see Table 1).

Independent validation was provided using two additional, external and independent, datasets reserved strictly for validation: the external validation (EVA) is on 6962 records and the independent validation (IVA) is on 2878 records. The independent datasets used for validation were collected in different medical centers with different cardiologists from the training dataset.

### 4.2 ECG findings versus HART-Findings

ECG’s limited diagnostic capability for the mentioned HF related structural abnormalities is confirmed by the literature, as is summarized in section 2.2.1. All too often, the ECG is nondiagnostic (inconclusive), and echocardiography confirmation is required. For example, patients with a classic description of anginal chest pain may have normal or non-specifically abnormal ECG, but evaluation of wall motion by echocardiography makes the diagnosis clear [43][44].

HART-findings diagnostic capability for HF related ECHO-findings is demonstrated by its higher diagnostic performance compared to the standard ECG interpretation. This comparison is shown in Table 2 by 8 ECHO-findings, and the results confirm that the Cardio-HART solution is better when compared to the best ECG-findings.

**Table 2.**
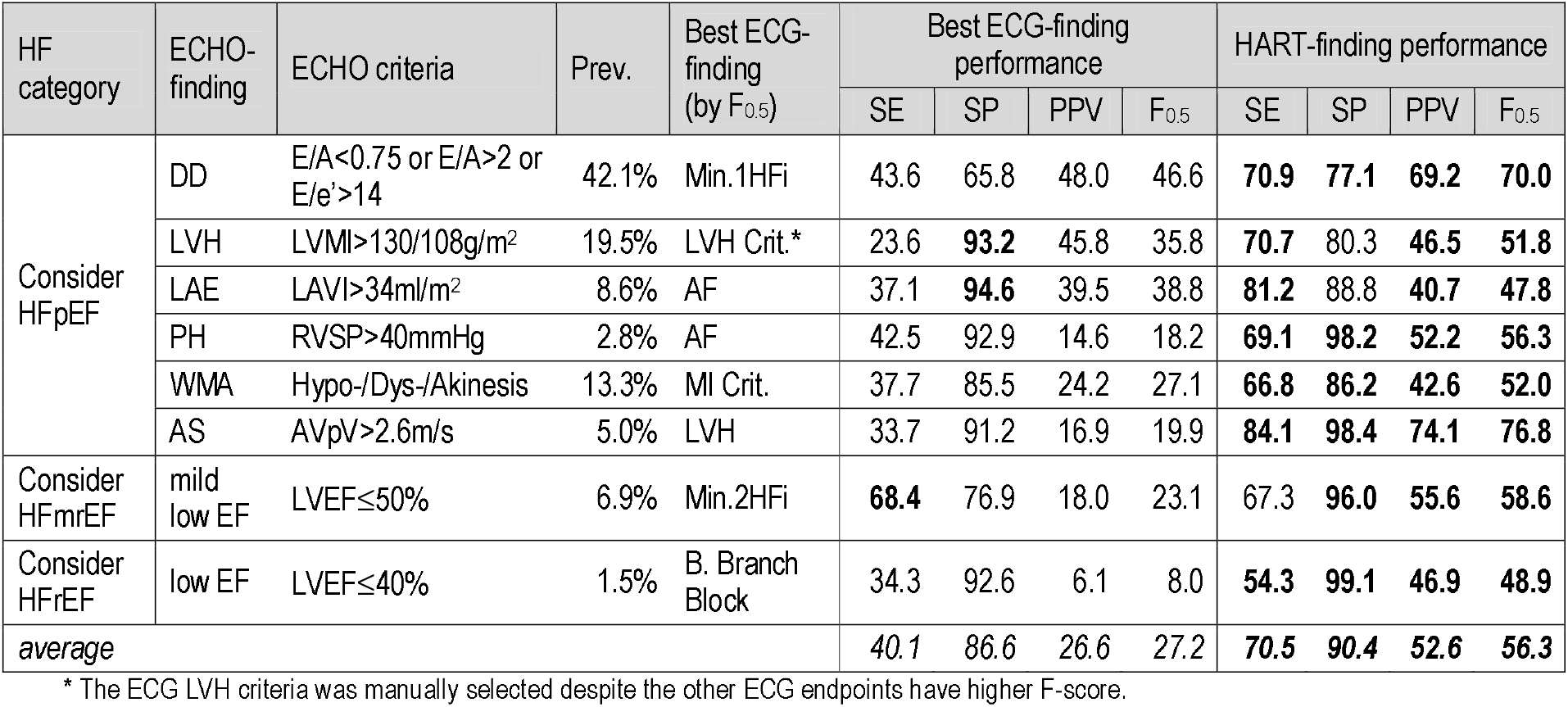
List of HF related ECHO-findings and diagnostic performance of the best ECG-findings and HART-findings.

The most important and preselected rule-based ECG-findings are evaluated in this comparison:

- Atrial Fibrillation (AF)
- Leftward Axis (LAX)
- Bundle Branch Block (BBB) including both Left and Right BBB
- ST-T deviation (ST-T) when ST interval and T abnormal at the same time, but excluding non-specific ST deviation
- T-wave abnormalities (TWA), including ischemic T-wave inversion,
- Atrial Abnormality ECG criteria (AA Crit.), including both Left and Right Atrial Abnormality
- LV Hypertrophy ECG criteria (LVH Crit.)
- Ventricular Hypertrophy (VH Crit.), including both LVH and RVH criteria
- Myocardial Infarction ECG criteria (MI Crit.) including STEMI and pathological Q wave criteria
- Minimum 3 HF indicator (Min.3HFi): minimum 3 abnormality occurs together from the following 9 HF indicator findings: AF, LAX, BBB, ST-T, TWA, AA Crit., VH Crit., MI Crit., or Tachycardia
- Minimum 2 HF indicator (Min.2HFi): minimum 2 abnormality occurs together from the 9 HF indicator findings.
- Minimum 1 HF indicator (Min.1HFi): minimum 1 abnormality occurs together from the 9 HF indicator findings.

The top graph of Fig. 4 shows the F^0.5^-score performance comparison of all the examined ECG-findings and HART-findings. The two bottom graphs compare the best ECG and HART-Findings in ROC and PRC. The area under the curve (AUC) is estimated based on the average sensitivity and specificity, which show a significant increase from ECG to HART as listed in Table 2: HART has higher sensitivity, specificity, and PPV, almost double the F^0.5^-score.

**Figure 4.**
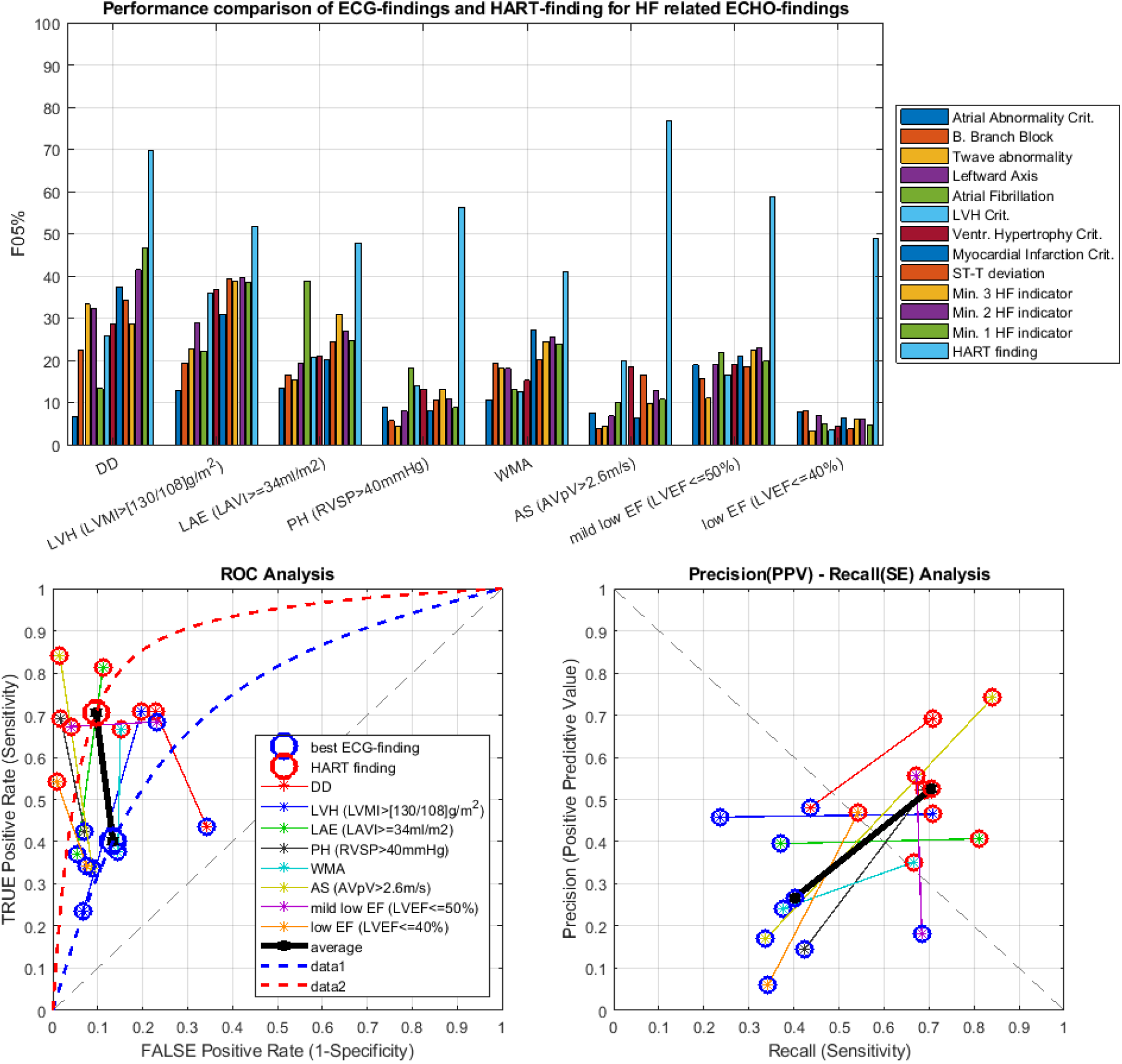
Performance comparison of ECG-findings and HART-findings for HF related ECHO-findings. (A) F-score the overall prediction capability of the examined findings for ECHO-findings and average (B) ROC comparison between ECG and HART, C) PRC comparison between ECG and HART

HART-Findings resolve half of the FP or FN, and uncertainties in the detection of HF-related abnormalities compared to ECG findings, a breakthrough for the non-invasive and bio-signal-based HF screening task.

### 4.3 HF performance by HART-findings

The prediction of HF categories by the HART-findings is evaluated with the help of ground truth HF categories. To estimate the performance, validation datasets were used as mentioned in section 4.1.

Table 3 shows the HF quad-format confusion matrix for an external (EVA) and independent validation (IVA) dataset, and HF binary performance on these two datasets is compared to ECG summary performance. The joined validation (EVA+IVA) database is a multi-center database, which allows for enhancement of reproducibility and generalizability [45].

**Table 3.**
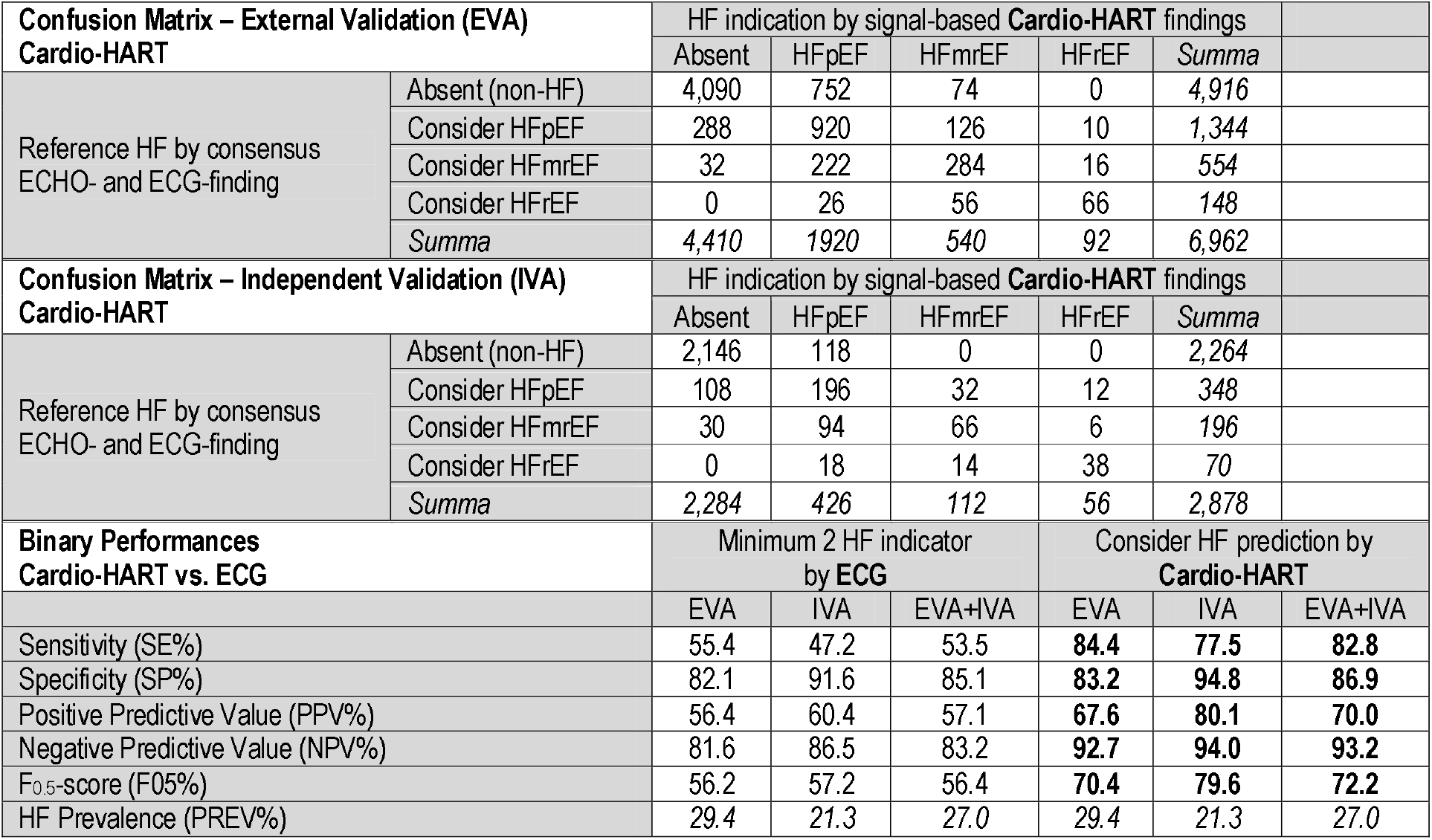
Confusion matrix on HF classification by the Cardio-HART system and Binary Performance Evaluation of HF classification by Cardio-HART system compared to ECG’s best HF criteria (Min.2HFi)

| Confusion Matrix – External Validation (EVA)<br>Cardio-HART |  | HF indication by signal-based Cardio-HART findings |  |  |  |  |  |
| --- | --- | --- | --- | --- | --- | --- | --- |
|  |  | Absent | HFpEF | HFmrEF | HFrEF | Summa |  |
| Reference HF by consensus<br>ECHO- and ECG-finding | Absent (non-HF) | 4,090 | 752 | 74 | 0 | 4,916 |  |
|  | Consider HFpEF | 288 | 920 | 126 | 10 | 1,344 |  |
|  | Consider HFmrEF | 32 | 222 | 284 | 16 | 554 |  |
|  | Consider HFrEF | 0 | 26 | 56 | 66 | 148 |  |
|  | Summa | 4,410 | 1920 | 540 | 92 | 6,962 |  |
| Confusion Matrix – Independent Validation (IVA)<br>Cardio-HART |  | HF indication by signal-based Cardio-HART findings |  |  |  |  |  |
|  |  | Absent | HFpEF | HFmrEF | HFrEF | Summa |  |
| Reference HF by consensus<br>ECHO- and ECG-finding | Absent (non-HF) | 2,146 | 118 | 0 | 0 | 2,264 |  |
|  | Consider HFpEF | 108 | 196 | 32 | 12 | 348 |  |
|  | Consider HFmrEF | 30 | 94 | 66 | 6 | 196 |  |
|  | Consider HFrEF | 0 | 18 | 14 | 38 | 70 |  |
|  | Summa | 2,284 | 426 | 112 | 56 | 2,878 |  |
| Binary Performances<br>Cardio-HART vs. ECG |  | Minimum 2 HF indicator<br>by ECG |  |  | Consider HF prediction by<br>Cardio-HART |  |  |
|  |  | EVA | IVA | EVA+IVA | EVA | IVA | EVA+IVA |
| Sensitivity (SE%) |  | 55.4 | 47.2 | 53.5 | 84.4 | 77.5 | 82.8 |
| Specificity (SP%) |  | 82.1 | 91.6 | 85.1 | 83.2 | 94.8 | 86.9 |
| Positive Predictive Value (PPV%) |  | 56.4 | 60.4 | 57.1 | 67.6 | 80.1 | 70.0 |
| Negative Predictive Value (NPV%) |  | 81.6 | 86.5 | 83.2 | 92.7 | 94.0 | 93.2 |
| $F_{0.5}$-score ($F_{0.5}$%) | | 56.2 | 57.2 | 56.4 | 70.4 | 79.6 | 72.2 |
| HF Prevalence (PREV%) |  | 29.4 | 21.3 | 27.0 | 29.4 | 21.3 | 27.0 |

Cardio-HART reached higher performance in prediction of HF when compared to the best ECG-based criteria: sensitivity increased from 53.5% to 82.8%, specificity from 85.1% to 86.9%, positive predictive value from 57.1% to 70.0%, and the F^0.5^-score from 56.4% to 72.2%. - calculated for combined validation sets, see performance under EVA+IVA in Table 3.

Fig. 5 shows the parametric ROC and PRC comparison of HF prediction between selected ECG-findings and the Cardio-HART (CHART) HF model, which is based on its own signal-based HART-findings. On the sensitivity/1-specificity plane (ROC), the individual ECG-findings have conspicuously low sensitivity from 5 to 35%, such as Atrial Fibrillation, ST-T deviation, MI, or B. Branch Block. The Minimum 1 HF indicator ECG criteria shows lower specificity, which means high false positives in the real-world situation. A more acceptable low false positive rate was provided by the Minimum 2 HF indicator ECG criteria, which reached 53.5% sensitivity (evaluated in Table 3). The minimum 3 HF indicator has a high specificity of 93% but reaches only poor sensitivity of 31%. The PRC confirms the ROC results but complements them with PPV, which makes it more visible, showing that both the PPV and sensitivity are increased by Cardio-HART compared to the best ECG criteria.

**Figure 5.**
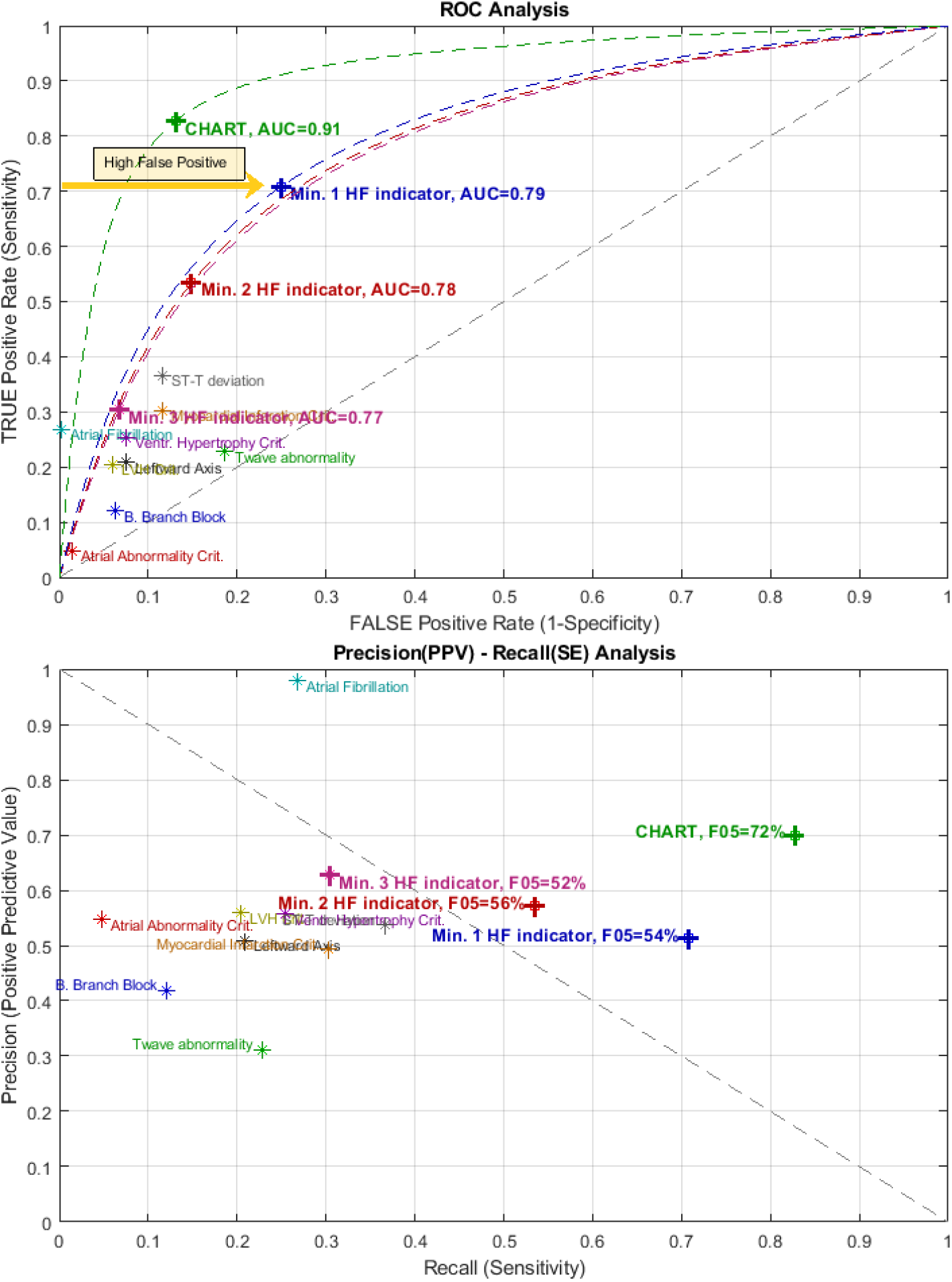
ROC and PRC analysis of Cardio-HART (CHART) and various ECG criteria for HF detection.

## 5 Discussion

The HFmrEF and HFrEF categories of HF can be successfully detected with models that predict low LVEF or simply LVSD. In contrast, HFpEF is a complex and diversified condition that cannot be classified as only diastolic HF or as an abnormal ECG patient. HFpEF detection relies on simultaneous abnormalities such as arrhythmias, structural heart diseases (LVH, LAE), valve diseases (most prevalent are AR, AS, MR, TR) and diastolic dysfunction. HART-findings can effectively detect these underlying conditions using multiple but synchronized bio-signals that can define HF.

### 5.1 Implications

HFpEF is where the greatest beneficial treatments can be made to improve HF [3][46]. Although studies have shown that the death rate for HFpEF is comparable to HFmrEF, it is lower than HFrEF [47][48]. Yet, HFpEF accounts for half of all hospitalizations of patients over 65 [49], posing a significant burden to healthcare.

The implications of this breakthrough technology will resonate throughout the healthcare system starting in Primary Care, where this low-cost, non-invasive, immediate use on initial patient presentation will, 1) drastically reduce time to diagnosis from months, to one day; 2) enable early detection of HF, by phenotype; which in turn 3) provide access to treatments earlier, 4) reduce the damage caused by delayed diagnosis; 5) reduce failures to diagnose, especially in women patients [50]; and, 6) clinically proven to reduce unnecessary referrals to cardiology [51]. The potential costs savings to healthcare are significant.

The impact also accrues to cardiology; they will get a better patient. As a direct substitute for ECG, prior to conducting a full echo examination, the implications are [51]: 1) screening for true negative patients that don’t need an echo; 2) screening for the presence of HF, valve disease, and myocardial infarctions, when the incoming referral fails to indicate these conditions; and critically, 3) providing a “big picture” of a patient’s overall cardiac status and revealing potential comorbidities that might otherwise be missed.

Cardio-HART is not a substitute for echocardiography when echocardiography is indicated. However, where echocardiography might not be available, CHART can provide substantially equivalent insight into the patient’s condition that can help determine the patient’s pathway and its priority based on urgency and severity. Various uses in secondary care include: 1) pre-surgery to determine if the patient has a cardiac abnormality, in particular HF, that might put the patient at risk during surgery; 2) respiratory care, where COPD and CVD often exist together; 3) Oncology, for early diagnosis of radiation induced heart disease, especially HF; 4) Emergency, after stabilization, confirm first if heart related or not, and then determine appropriate patient pathway; and 5) cardiology in general, where patient rounds could be done on the basis of ECHO findings, including HF, and not merely ECG.

Abnormalities associated with the right heart side are indicative of HFrEF, especially right atrial and ventricular enlargement. The bio-signal diagnostic capability for the right side of the heart has already been indicated by various research, e.g., TR prediction through ECG RAE criteria and PCG/MCG murmurs [59] or analysis of second heart sound splitting in evaluation of PH [60]. Compared to ECG RVH criteria, HART RAE and HART RVE findings are far more sensitive criteria, relevant for COPD patients, in particular those at risk of HF.

In all cases, CHART AI, in the form of HART findings, provides for a more immediate determination of a patient’s cardiac status and access to treatment.

### 5.2 Conclusion

Detection of HF should be on initial patient presentation to primary care, provided by non-cardiologists [3]. The standard ECG has limitations and is not sufficiently sensitive or specific for physiological cardiac dysfunction and, as a result, generates many false positives. There is an urgent, unmet medical need in primary care for diagnosing HF, in particular the heart structural, functional, and hemodynamic abnormalities that define the specific condition, as this would allow for the immediate start of treatment as per the guidelines, thereby stopping disease progression. In a world of backlogs and long wait-times, this is critical to better patient outcomes.

HART-findings have been shown to address this unmet medical need. Cardio-Phoenix’s Cardio-HART solution provides an understanding of both left heart side abnormalities (LVH, LAE, MR, and WMA) and right heart side abnormalities (RVE, RAE, TR, and PH) and can detect the origin and severity of HF. That enhances diagnosis, risk stratification, and prioritization for patients. Cardio-HART HF prediction reaches significantly higher overall performance compared to the best ECG criteria, with sensitivity of 83%, specificity of 87% and a positive predictive value of 70%. Higher sensitivity means lower false negatives and early detection. Similar specificity means a not higher rate of false positives, typically a major problem for CNN-like solutions.

These clinically meaningful results are thanks to the fact that Cardio-HART’s complex machine learning system processes not only ECG signals but also two other bio-signals, PCG and MCG, that together combine to help provide a multi-dimensional perspective of cardiac dysfunctions critical in identifying HF and other major CHART findings. Significantly, Cardio-HART’s AI strategy is based on learning networks that incorporate the last 50 years of knowledge about these bio-signals and cardiac dysfunctions, which we call Knowledge-enhanced Neural Networks.

## Data Availability

Data are available on reasonable request. The clinical data are for legitimate purposes on request. The Official Study Report can be obtained on request

## 6 Acknowledgments

The collection of the data used in this paper was previously collected in clinical investigations funded in part by UVA Research Corp. No other outside funding was involved.

## 8 Appendices

### 8.1 Abbreviations

ECG: Electrocardiography
ECHO: Echocardiography
PCG: Phonocardiography
MCG: Mechanical force bio-signal
CHART: Cardio-HART™ from Cardio-Phoenix
HART: ECHO-like findings estimated by Cardio-HART AI system
HF: Heart Failure
HFpEF: HF with preserved ejection fraction
HFmrEF: HF with mildly reduced ejection fraction
HFrEF: HF with reduced ejection fraction
LSTM: Long short-term memory network
HHT: Hilbert–Huang transform
PSD: Windowed Power Spectrum Density
MFCC: Mel-frequency cepstral coefficients
JMIM: Joint Mutual Information Maximization
JRMI: Renyi entropy-based Joint Mutual Information
ECFS: Eigenvector Centrality Feature Selection
FSMB: Markov blanket-based Feature Selection
KENN: Knowledge-enhanced neural networks
BNP: Natriuretic peptides
LVSD: Left Ventricular Systolic Dysfunction
ALVSD: Asymptomatic LVSD
DD: Diastolic Dysfunction
DDIM: Impaired Relaxation type DD
LVH: Left Ventricular Hypertrophy
RVH: Right Ventricular Hypertrophy
DCM: Dilated Cardiomyopathy
RVE: Right Ventricular Enlargement
LAE: Left Atrial Enlargement
RAE: Right Atrial Enlargement
WMA: Wall Motion Abnormality
AS: Aortic Stenosis
MS: Mitral Stenosis
MR: Mitral Regurgitation
AR: Aortic Regurgitation
TR: Tricuspid regurgitation
PH: Pulmonary Hypertension
CAD: Coronary Artery Disease
AF: Atrial Fibrillation
LAX: Leftward Axis
BBB: Bundle Brach Block
ST-T: ST interval and T abnormal
MI: Myocardial Infarction
LVH: LV Hypertrophy
RVH: RV Hypertrophy
LVEF: Left Ventricular Ejection Fraction [%]
GLS: Global longitudinal strain
SV: Stroke Volume
FP: False positive
FN: False negative
SE: Sensitivity
SP: Specificity
PPV: Positive Predictive Value
F05: F-score with β=0.5
AVpV: Aortic Valve Peak Velocity [m/s]
MV-E: Mitral Inflow E Wave Velocity [m/s]
E/A: E/A Wave Velocity on Mitral Valve [m/s/m/s]
LVMI: Left Ventricular Mass Index [g/m^2^]
LVIDd: End-diastolic Left Ventricular Diameter [mm]
LVIDs: Endsystolic Left Ventricular Diameter [mm]
LAVI: Left Atrial Volume Index [mL/m^2^]
RAVI: Right Atrial Volume Index [mL/m^2^]
RVSP: Right Ventricular Systolic Pressure [mmHg]
MRj/LA: Mitral Regurgitation Jet Ratio in Left Atrium [%]
TRj/RA: Tricuspid Regurgitation Jet Ratio in Right Atrium [%]

#### 8.2 Declaration of Helsinki

All patients were recruited in accordance with the Helsinki protocol and each provided written informed consent to participate.

#### 8.3 Copyrights

All copyrights and trademarks are owned by their respective owners, and all rights are attributed to them. Cardio-Phoenix, Cardio-HART, C.H.A.R.T, Cardio-TriTest, are trademarks of Cardio-Phoenix Inc. (all rights are reserved worldwide).

#### 8.4 Author Contributors

- Consultant cardiologists in research: Marta Afonso Nogueira, Simone Calcagno
- Final approval of the article: Niall Campbell, Georgios Koulaouzidis
- Literature research: Azfar Zaman, Istvan Kecskes
- Acquisition of data and ground truth definition: Tatjana Stankovic, Erzsebet Szabo, Aniko B. Szabo
- Data verification: Anwar Jalil, Firdous Alam
- Statistical analysis and administration: Istvan Kecskes

All authors have approved the final article.

#### 8.5 Graphical Abstract

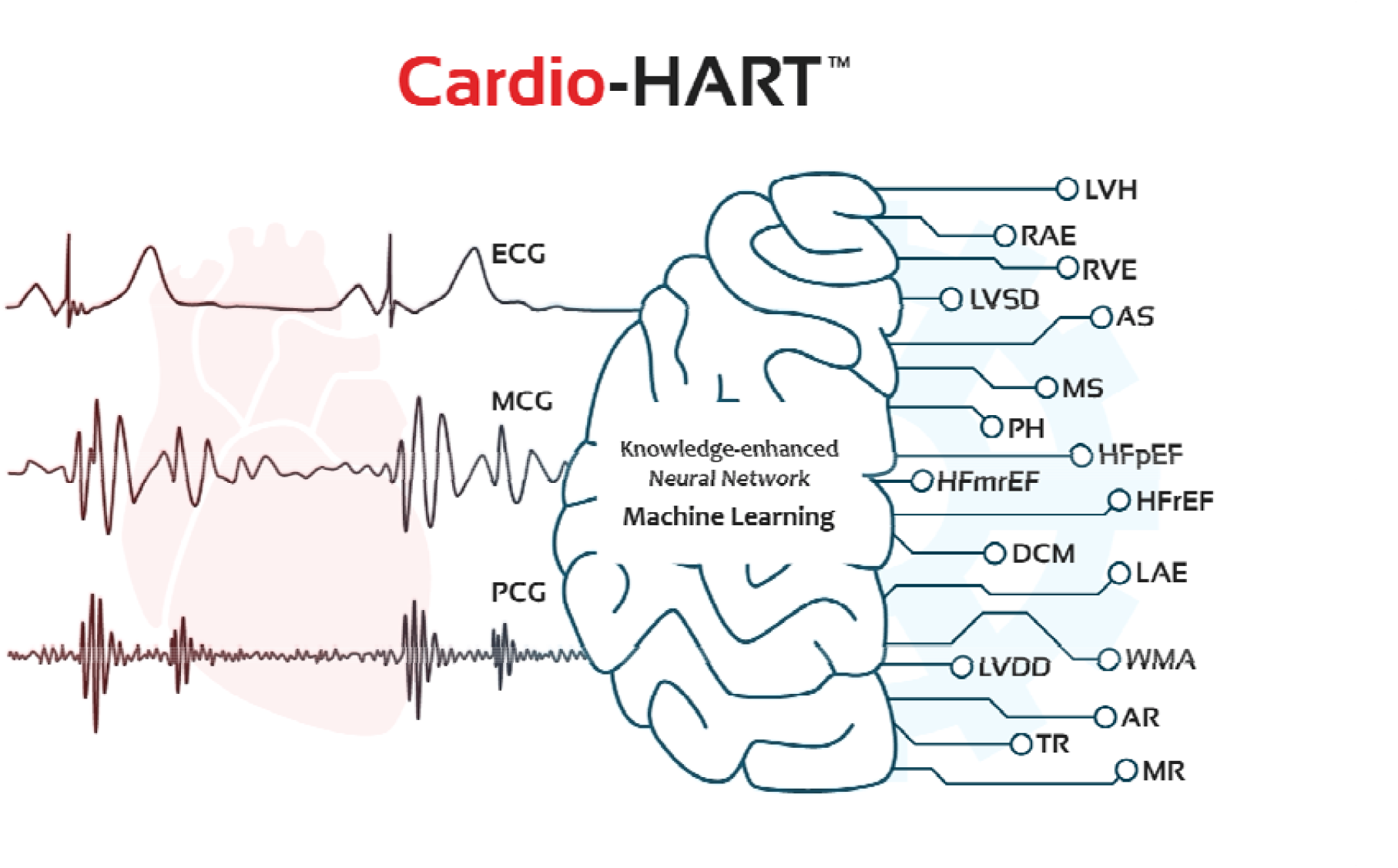

## Footnotes

1 FDA requested that HART findings be used instead of Echo-findings to better indicate their bio-signal origin.

2 multi-parameters mean more than one measurement is considered or required in the practical application of

3 https://www.cardiophoenix.com/

